# The timing and effectiveness of implementing mild interventions of COVID-19 in large industrial regions via a synthetic control method

**DOI:** 10.1101/2020.06.22.20137380

**Authors:** Ting Tian, Wenxiang Luo, Jianbin Tan, Yukang Jiang, Minqiong Chen, Wenliang Pan, Songpan Yang, Jiashu Zhao, Xueqin Wang, Heping Zhang

## Abstract

The outbreak of novel coronavirus disease (COVID-19) has spread around the world since it was detected in December 2019. The Chinese government executed a series of interventions to curb the pandemic. The “battle” against COVID-19 in Shenzhen, China is valuable because populated industrial cities are the epic centres of COVID-19 in many regions. We made use of synthetic control methods to create a reference population matching specific characteristics of Shenzhen. With both the synthetic and observed data, we introduced an epidemic compartmental model to compare the spread of COVID-19 between Shenzhen and its counterpart regions in the United States that didn’t implement interventions for policy evaluation. Once the effects of policy interventions adopted in Shenzhen were estimated, the delay effects of those interventions were referred to provide the further control degree of interventions. Thus, the hypothetical epidemic situations in Shenzhen were inferred by using time-varying reproduction numbers in the proposed SIHR (Susceptible, Infectious, Hospitalized, Removed) model and considering if the interventions were delayed by 0 day to 5 days. The expected cumulative confirmed cases would be 1546, which is 5.75 times of the observed cumulative confirmed cases of 269 in Shenzhen on February 3, 2020, based on the data from the counterpart counties (mainly from Broward, New York, Santa Clara, Pinellas, and Westchester) in the United States. If the interventions were delayed by 5 days from the day when the interventions started, the expected cumulative confirmed cases of COVID-19 in Shenzhen on February 3, 2020 would be 676 with 95% credible interval (303,1959). Early implementation of mild interventions can subdue the epidemic of COVID-19. The later the interventions were implemented, the more severe the epidemic was in the hard-hit areas. Mild interventions are less damaging to the society but can be effective when implemented early.

AMS 2000 subject classifications: Primary 00K00, 00K01; secondary 00K02.

## 1. INTRODUCTION

The confirmed cases of novel coronavirus (SARS-Cov-2) disease (COVID-19) had been detected since December 2019[30], and there are over 200 countries with reported confirmed cases in the world[35]. A potent characteristic of SARS-Cov-2 is that it can be contagious during the incubation period, where asymptomatic individuals could become a disseminating source[28]. Therefore, finding and implementing effective interventions is vital to disease control[17, 32].

On January 23, 2020, non-pharmaceutical interventions including all public transportation closures started in Wuhan, and a similar response was triggered in cities geographically near Wuhan cities, such as the cities within Hubei province and Wenzhou in Zhejiang province on February 1, 2020[20]. Guangdong province, where there was a large population of migrants from Hubei province[8], activated the first-level public health emergency response on January 23, 2020[34]. Shenzhen, with confirmed cases of COVID-19 initially reported in Guangdong province, implemented the interventions by closely following and isolating the close contacts of confirmed cases of COVID-19. By far, the strategic polices for direct protection (such as wearing face masks) and for transmission reduction (for example 14 days isolation for oversea travellers, cancellation of public gatherings, delayed reopening of schools) have been maintained in Shenzhen. The intervention and control strategies of Shenzhen were adopted as a classical example in the report from World Health Organization (WHO)[5].

As an industrial city, where there was a large population of inbound and outbound travellers[43], Shenzhen implemented a relatively mild intervention strategy compared with that of Wuhan and mounted an early response to the epidemic of COVID-19 relative to other cities such as Wenzhou. Therefore, there is a great interest to examine how effective such mild but early interventions were in curbing the epidemic of COVID-19 in Shenzhen. We evaluated the treatment effects of intervention policies in Shenzhen by comparing the epidemic data in Shenzhen with its counterpart regions in the United States counties, and simulate the potential outcomes of different hypothetical starting dates of interventions to project the influence of the delay effects on the policy effectiveness.

To compare the effects of the control of epidemic in Shenzhen, the synthetic control method[2] as a classical methodology of causal inference was modified. It estimated the effects of short-term interventions between the affected unit and a combination of unaffected units[1]. To seek a series of unaffected units, i.e. the regions where interventions similar to those adopted in Shenzhen were not issued at the early stage of epidemic, we employed the infected counties in the United States as the quantitative reference for comparison.

Alternatively, in order to evaluate the delay effects of policy interventions appropriately, it’s crucial to consider the pre-symptomatic transmission of COVID-19[16] to reflect the nature of COVID-19. Traditional epidemic models, such as SIR (Susceptible, Infectious, and Recovered) mode[14, 11] and SEIR (Susceptible, Exposed, Infectious, and Removed) model[29, 26], may not be suitable for modelling the spread of COVID-19[42] due to the lack of consideration for the incubation period and the pre-symptomatic transmission pattern during the incubation period. The literature suggests that the mean length of the incubation period ranges from 5 to 7 days, and considering the incubation period is important to the understanding of disease risk[28, 30, 7, 31].

In this paper, we proposed the SIHR (Susceptible, Infectious, Hospitalized, Removed) model, which considers pre-symptomatic transmission of COVID-19 by an appropriate compartmental assumption with the well-defined time-varying reproduction number. By modifying the estimated time-varying reproduction number[24, 36] to reflect delay effects of interventions, we simulated different outcomes of COVID-19 for comparison.

The rest of the paper is organized as follows. Section 2 describes the synthetic control method with a modified selection of control variables and the proposed SIHR model. The results of treatment effects and delay effects of mild interventions are reported in Section 3. We end this paper with some concluding remarks.

## 2. METHODS

### 2.1 Data Collection

Epidemic data were collected for daily cumulative confirmed cases of COVID-19 since January 19, 2020 in Shenzhen when the first confirmed case was reported[44]. Its corresponding population, latitude and areas were collected in the statistical yearbook. Also, the daily cumulative confirmed cases of COVID-19 for each county of each state in the United States were downloaded and available online[18] since March 1, 2020. The corresponding populations, areas, and latitudes were collected from the United States Census Bureau (USCB)[12, 13].

### 2.2 Effect of mild interventions

To examine the treatment effects of interventions implemented in Shenzhen, we envisioned that cumulative numbers of confirmed cases of COVID-19 per 100,000 persons in the population could be observed at the time *t* whether the interventions were implemented (treated) or not (not treated), which are denoted by 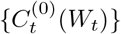.

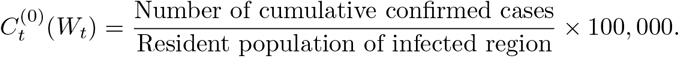

Here *t* is the time and *W_t_* is the group indicator. “*W_t_* = 1” corresponds to “exposed to interventions”, and “*W_t_* = 0” otherwise. Thus, the potential outcomes at the time *t* are 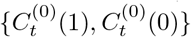 and the corresponding treatment effect of interventions is 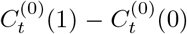. However, only one of the potential outcomes could be observed, i.e. the observed outcome at time *t* is defined as:

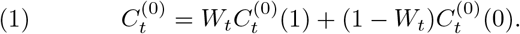

Here the untreated outcomes 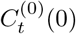 of “absence of interventions” were missing, which are the so-called counter-factual outcomes[38]. To formalize 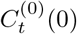, we modified the synthetic control method (SCM)[2, 1] to construct the counterfactual outcomes by combining a number of untreated units instead of a single untreated unit.

Those potential untreated units were selected from the United States counties, where the interventions were not adopted at the early stage of epidemic. The epidemic data were reported on March 1, 2020 in the most infected counties of the United States[18]. However, the White House issued a “call to action” for coronavirus guidelines including cancelling gatherings over 10 people and staying at home starting on March 16, 2020[22]. There were no broad interventions implemented in the early stage of the epidemic in the United States, which allowed us to define a pre-intervention period from March 1 to March 16. Therefore, there were 16 days of duration before the interventions implemented in the United States. These 16-day epidemic data in the United States were used as the benchmark in our comparison with the 16-day epidemic data in Shenzhen from January 19 to February 3, 2020.

COVID-19 was detected and initially reported on January 19, 2020 in Shenzhen[44], and Guangdong province that includes Shenzhen as one of its cities activated “public responses” on January 23, 2020[34], so there were 4 days before Shenzhen implemented the interventions after the first case was reported. We use *T*_0_ = 4 to denote this preintervention period, which is 4. We compared the data between Shenzhen from January 19 to February 3, 2020 and counties in the United States from March 1 to March 16, 2020, by fixing the total duration at 16 days starting from the day of the first reported case in the locations of interest, i.e. “*t* = 1” means January 19 for Shenzhen, and March 1 for the counties of the United States. The effect of the postintervention period at time *t* is 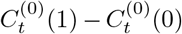 for *t* > *T*_0_. It can be estimated by:

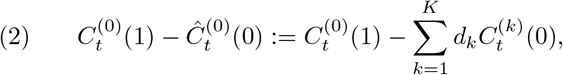

where 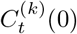 was the observed cumulative numbers of COVID-19 confirmed cases per 100,000 at the time *t* in *k*-th county of the United States in the absence of interventions. Thus, the counterfactual outcomes in the absence of the interventions of Shenzhen during the post-intervention period were calculated from a weighted linear combination of *K* untreated counties. The weights, *d_k_*’s, were non-negative and summed to 1[1]. Reliable estimates, 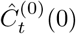, can be obtained by constructing a synthetic region whose characteristics was similar to that in Shenzhen in the pre-intervention period[3].

The strategy is to find the important characteristics between Shenzhen and its “highly resembling” regions relating to the transmission of COVID-19 but not affected by the interventions. The literature suggests that the risk of transmission is higher in denser populations, and more urbanized regions may have more infected individuals[25, 15]. There is a large number of infected counties in the United States, and the population density of each county varies dramatically from near 0 to 18,721 persons per square kilometre. We selected the counties with the sizes of population densities (*P*) close to Shenzhen, i.e. metropolis areas in the United States.

We also considered latitude (*L*) of the counties because the latitude was found to be associated with the transmission of the COVID-19[39, 25, 40], before applying the data assimilation[37] procedure in the construction of the synthetic region.

Let 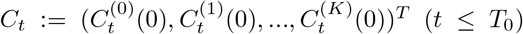. In order to estimate *d_k_* appropriately, we assumed that *P* and *L* share a common least square structure related to SCM[1, 2]:

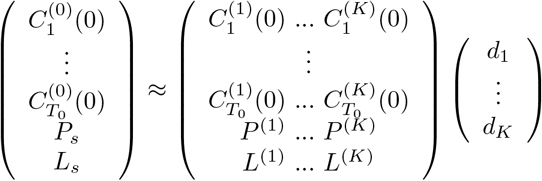

where *P_s_* and *L_s_* are the population density and the latitude of Shenzhen, respectively. Similarly, *P*^(^*^k^*^)^ and *L*^(^*^k^*^)^ are the population density and the latitude of the *k*-th county of the United States, respectively.

The literature[2, 1] recommended that not all the *C*_t_, *t* ≤ *T*_0_ should be included in the estimation procedure, although the user can choose to include as many as *M* (*M* ≤ *T*_0_) linearly independent combinations of pre-intervention outcomes [2]. Let

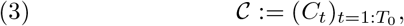

which can be viewed as the spatial-temporal data for our data. We performed principal component analysis (PCA)[19, 23] for the columns of 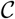 and evaluated the value of *M* and the linearly independent combinations of pre-intervention outcomes. The PCA reduced the redundant information of time signal and the impact of cumulative confirmed cases within the estimation procedure.

Let 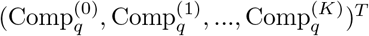 be the *q*-th component of the columns of 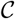 in the PCA. Also, let 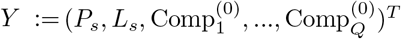 be a (J × 1) vector, where *J* = *Q* + 2, and *Q* is the chosen number of components. In other words, *Q*: = *M*. We can choose *Q* to be the smallest number of the ranked principal components that explain at least 95% of the variance in 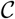. *X* is a (*J* ×*K*) matrix whose *k*-th column is 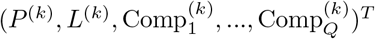. The vector *D*: = (*d*_1_,…, *d_K_*)*^T^* is obtained by minimizing the distance 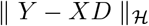:

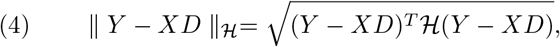

where 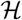 is a *J* × *J* positive semidefinite matrix. The estimated weight 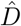 was obtained by Nelder-Mead methods[33, 27].

In addition to the population density and latitude, there are other factors related to the transmission of COVID-19. However, there were only 4 days in the pre-interventions period, the inclusion of more predictors in the linear construction of “synthetic Shenzhen” would result in collinearity during this short pre-intervention period; for example, temperature and latitude are correlated with each other. Alternatively, the *Q* components of pre-intervention outcomes included in the construction of “synthetic Shenzhen” can reflect the unobserved, shared, and time varying factors[3]. The gaps in the health care system might contribute to the variation in the number of confirmed cases in infected regions. However, the intervention and treatment of the COVID-19 depend on national and local authorities and resources. In the construction of “synthetic Shenzhen”, we focus on the natural characteristics that are not easily affected by human decisions.

After the synthetic region of Shenzhen was constructed, an inference for the treatment effect of interventions was performed by a placebo test[3]. A “placebo effect”[4] was obtained by assigning an arbitrary sample of K counties in the United States as a “treated unit”, Shenzhen and the remaining (*K* − 1) counties synthesized to a combined untreated unit, the difference (gap) of COVID-19 cases per 100,000 between any of the “treated unit” and corresponding combined untreated unit during post-intervention period formed to a permutation distribution. If the value of this gap from Shenzhen was extreme compared to the remaining gaps in the permutation distribution, the treatment effect of interventions implemented in Shenzhen was considered statistically significant [2].

### 2.3 The delay effects of mild interventions

Besides the determination of treatment effect of intervention in Shenzhen, the timeliness of policy implementation may also affect the effectiveness of intervention. We considered the likely outcomes of the interventions if they were delayed by different days by using a SIHR model [42], which divided the total population into four classes: *S* (Susceptible to infectious disease), *I* (Infected and infectious without isolation), *H* (Hospitalized in well-isolation), and *R* (Removed from *H* because of the recovery or death). Let *Z_t_: =* (*S_t_,I_t_, H_t_, R_t_*) be the random numbers of compartments *S*, *I*, *H, R* at time *t*, s.t. *S_t_ + I_t_ + H_t_ + R_t_ = N*, where *N* was the total number of population. We assumed (*Z_t_*)*_t_*_∈1:_*_T_* followed the hidden Markov structure:

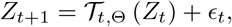

where 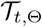 was the dynamic operator indexed by the time *t* and parameters Θ. *ϵ_t_* were some noise variables, which were independent at different times. We treated *I*_1:_*_T_* as the latent variables.

For the noise, we followed the literature in infectious disease [9] by assuming

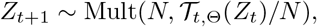

where Mult(*n*,*p*) was the density of the multinomial distribution with the total number *n* and the incident rate vector *p*. Based on the mechanism of the spread of epidemic, we assumed the following dynamical operator 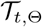[41]:

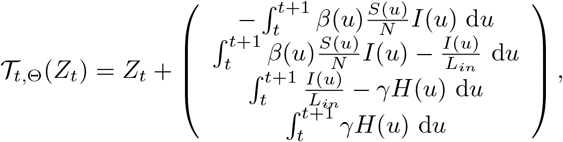

where (*S*(*t*), *I*(*t*), *H*(*t*), *R*(*t*)) = (*S_t_,I_t_*, *H_t_*, *R_t_*), and (*S*(*u*), *I*(*u*), *H*(*u*), *R*(*u*)) were determined by the dynamic system (A1) in the Appendix with the initial value (*S*(*t*), *I*(*t*), *H*(*t*), *R*(*t*)) (*t* < *u* ≤ *t* + 1). The functional parameter *β* was time-varying contact rate controlled by additional parameters *β*_0_, *m*_1_, *m*_2_. *L_in_* was the mean length of incubation period.

*L_in_* and γ can be assumed to be time-varying in SIHR model. However, since *I*_1:_*_T_* is unobserved, it may be crucial to fix *L_in_* to ensure identifiability, which was assumed to be 5 days for COVID-19[7, 21, 30]. Parameter γ corresponding to “the removed compartment” was considered in compartment *H* which did not affect the estimation of *β*(*t*). Because we focused on the SIHR model, we assumed γ to be time-invariant for simplicity.

Let Θ: = (*β*_0_, *m*_1_, m_2_, γ) are the parameters needed to be estimated.

To reflect the effect of interventions, we used the logistic functional] to simulate the decreasing trend of *β*(*t*):

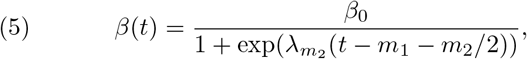

where 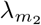 was chosen as 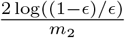 and *ϵ* was fixed to be 0.01. The smaller the values of *m*_1_ and *m*_2_, the earlier effectiveness and the stronger intensity of interventions were implemented, respectively[41]. The trend of *β*(*t*) was shown in Figure 1.

**Figure 1.**
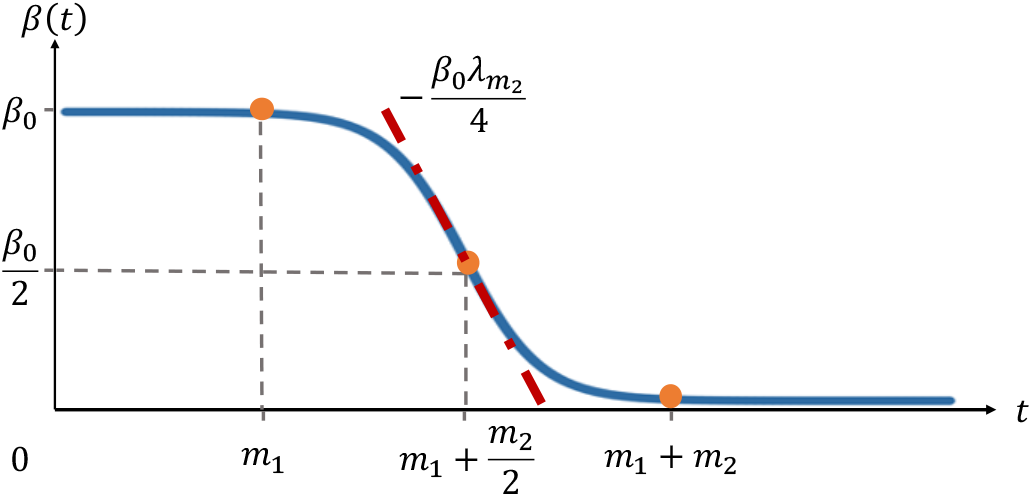
The trend of β(t)

The slope of *β*(*t*) at point 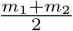 is 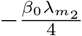. A larger value of 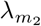, results in a larger slope, and the intensity of interventions would be stronger accordingly.

The time-varying reproduction number 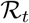[24, 36] of SIHR model was calculated from:

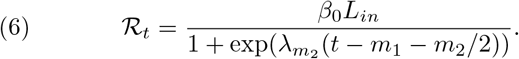

Given the observed data *H*_1:_*_T_* and *R*_1:_*_T_*, we used the posterior distribution:

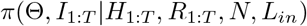

for Bayesian inference. Once the prior distribution of Θ: π(Θ) was given:

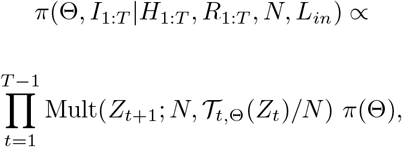

which could be randomly approximated by Gibbs sampler embedded Metropolis-Hastings steps [10, 6] since π(Θ, *I*_1:_*_T_*|*H*_1:_*_T_*, *R*_1:_*_T_*, *N*, *L_in_*) has no closed form. The full conditional distributions for the Gibbs sampler were:

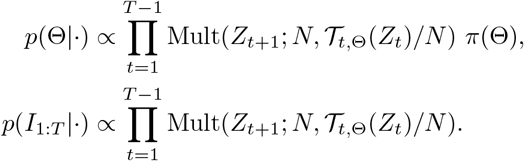

The initial values for Gibbs sampling were chosen as: Θ^(0)^ = (1,1,1,1) and 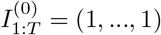. After a burn-in period in the Markov chain, we used the remained samples to approximate the posterior distribution[41, 42].

For the prediction of spread, we used the posterior distribution (*s* > *T*):

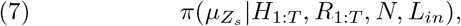

where *μZ_s_* was the conditional mean of *Z_s_* given *Z_T_*. Notice that:

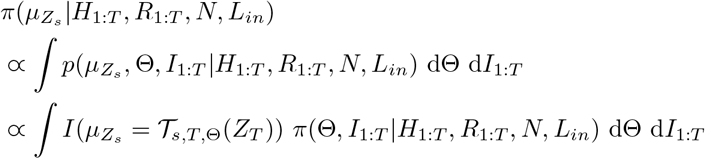

where 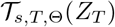 was the vector at time s determined by the dynamic system (A1) in the Appendix with the initial value *Z_T_* and parameters Θ. Hence the posterior distribution of was calculated from π(Θ,*I*_1:_*_T_*|*H*_1:_*_T_*, *R*_1:_*_T_*, *N*, *L_in_*).

The point estimation of parameters Θ, latent variables *I*_1:_*_T_* and the predicted 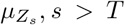, *s* > *T* were presented as the median of the posterior distribution while 95% credible intervals were constructed with 2.5% and 97.5% quantiles.

Parameter *m*_1_ reflected the effect of interventions over time. We varied the estimated values of *m*_1_ to investigate the likely outcomes of delayed interventions. We considered the delay operator 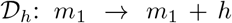 and let 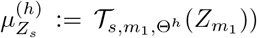, where 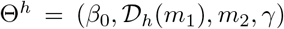, *s* > *m*_1_. Similarly, we used the posterior (7) of 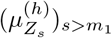 to simulate the potential outcomes of the delays of the interventions, where *h* varied from 0 to 5.

## 3. RESULTS

### 3.1 Synthetic control method (SCM) outcomes

Sixty-nine areas (68 counties in the United States and Shenzhen) were selected by using their corresponding population density over 500 people per square kilometre. SCM was used to combine 68 counties in the United States to construct a synthetic region of Shenzhen based on their latitude, population density and first two components from the PCA procedure described above, where the first two components explained over 95% of covariances of C in (3) for the pre-intervention period.

The synthetic and average values of latitude, population density, the first component, and the second component of 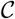 in (3) were shown in Table 1. During the pre-intervention period, the latitude was higher in the average of the 68 counties than in Shenzhen, while the population density was substantially lower on average in the 68 counties than in Shenzhen. Moreover, the values of two principal components of 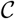 in (3) were negative on the average of the 68 counties. In contrast, the “synthetic Shenzhen” exactly reproduced the values of population density, and two principal components of 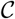 in (3) which had in Shenzhen. A “highly resembling” region of Shenzhen was obtained by mainly combining Broward (Florida), New York (New York), Santa Clara (California), Pinellas (Florida), and Westchester (New York) with their corresponding weights 0.423, 0.335, 0.123, 0.032 and 0.025 as an optimal solution, respectively. The weights of all possible counties (removed counties with value of weight 0) with their latitude and population density were represented in Table A1 in the Appendix.

**Table 1.** *The synthetic and average values of latitude, population density, the first component and the second component for 68 counties in the pre-intervention period*.

| Features | Shenzhen |  | Average of 68 counties |
| --- | --- | --- | --- |
|  | Real | Synthetic |  |
| Latitude | 22.55 | 33.16 | 38.15 |
| Population density | 6670 | 6669 | 1348 |
| The first component | 0.107 | 0.107 | -0.002 |
| The second component | 0.035 | 0.035 | -0.001 |

Before the mild intervention policies were implemented on January 23, 2020, the trends of actual cases per 100,000 in both Shenzhen and “synthetic Shenzhen” were highly similar, suggesting that such a synthetic region could be used to estimate the “counterfactual” results of Shenzhen. Based on Figure 2, after 1 day of the implementation of the policies, the growth rate of actual cases per 100,000 in Shenzhen was relatively slower than that of “synthetic Shenzhen” until January 29, 2020. After that, there was a sharp increase in the gap of cases per 100,000 between Shenzhen and “synthetic Shenzhen”. On the 16*^th^* day from January 19, 2020, i.e. February 3, 2020, the estimated number of cases per 100,000 in “synthetic Shenzhen” was 11.87, which is 5.75 times of the actual COVID-19 cases per 100,000 observed (2.07) in Shenzhen (Figure 2).

**Figure 2.**
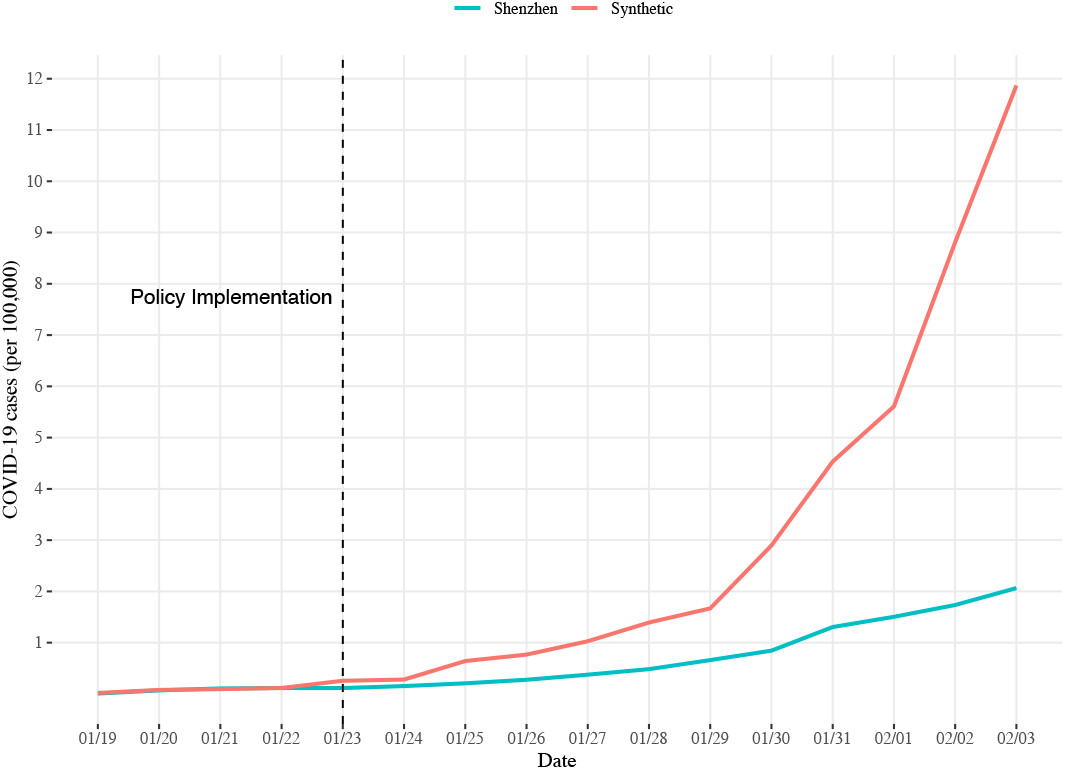
*The trends of COVID-19 cases per 100,000 between Shenzhen and “synthetic Shenzhen" from January 19 to February 3, 2020. The black dashed line indicated the day of starting the intervention policies in Shenzhen*.

A placebo test was performed to determine the significance level of the difference in the trends of COVID-19 cases per 100,000. To this end, we plotted the gap curves between Shenzhen and “synthetic Shenzhen” by in turn exchanging Shenzhen and each one of the 68 counties in the United States. The gap of COVID-19 cases per 100,000 between Shenzhen and our “synthetic Shenzhen” was the negatively largest among the negative gaps, i.e. the negative effect of the intervention on COVID-19 per 100,000 in Shenzhen was the lowest of all. For those 68 counties, the probability of having a gap for Shenzhen under a random permutation of the control measures was less than 5% (i.e. 1/69 = 0.014), which is statistically significant in a conventional test[2]. This suggested that the mild intervention policies of Shenzhen might have significantly reduced the COVID-19 cases per 100,000 (Figure 3).

**Figure 3.**
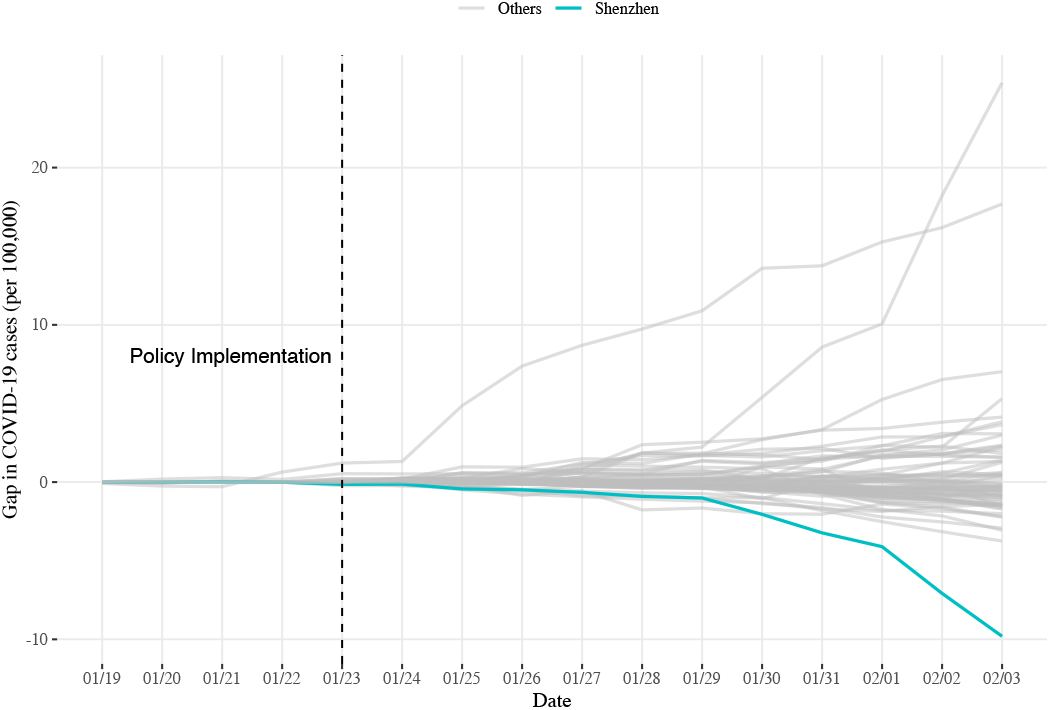
*The permutation test of the treatment effects of implementing the policies in Shenzhen and the 68 control regions in the United States. All grey curves represent the placebo tests of COVID-19 cases per 100,000 gaps between a control area* (*an arbitrary random county of the 68 counties*) *and synthetic control city* (*a combination of remaining 67 counties and Shenzhen*), *and the blue curve represents the placebo test of COVID-19 cases per 100,000 gap between Shenzhen and “synthetic Shenzhen"*.

### 3.2 The likely outcomes of delayed interventions

There were mild but early interventions implemented in Shenzhen. It is worthy examining the likely outcomes of those interventions if they were delayed by different days (Figure 4). The observed COVID-19 cases per 100,000 from January 19 to February 3, 2020 (red points in Figure 4) were very close to the 0-day delay curve (i.e. the estimated cumulative confirmed cases per 100,000). Based on our simulation by delaying the interventions, the expected number of cumulative confirmed cases per 100,000 would be 2.32 times for a 4-day delay and, a larger, 2.51 times for a 5-day delay on February 3, 2020 (Figure 4).

**Figure 4.**
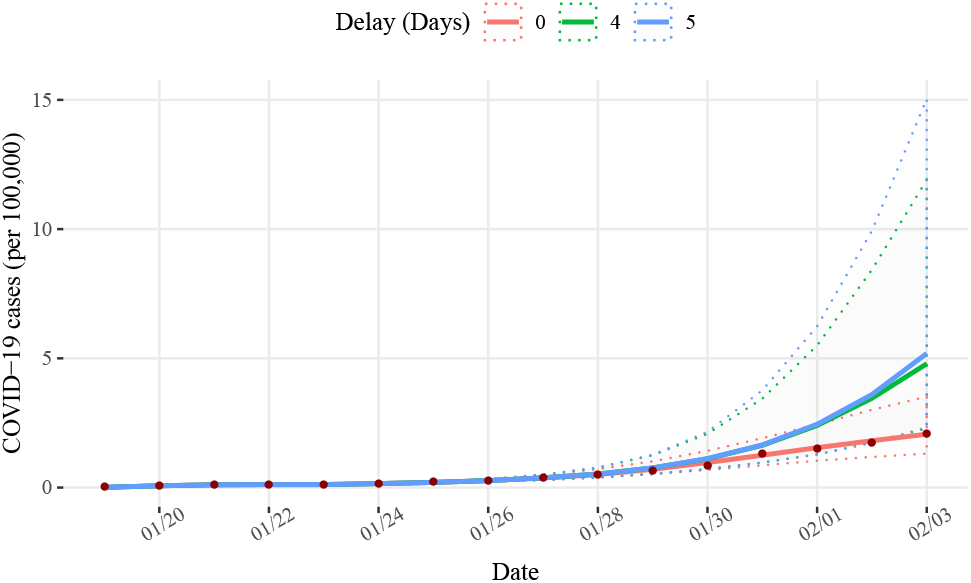
*The expected number of cumulative confirmed cases per 100,000 with corresponding confidence intervals for mild interventions if they were delayed by 0, 4 and 5 days. The points represent the actual cumulative confirmed cases per 100,000 of Shenzhen*.

### 3.3 Revisiting the results in terms of cumulative confirmed cases

We have presented our analysis and results based on the number of cumulative confirmed cases per 100,000 persons in the population. Here, we summarize our results based on the number of cumulative confirmed cases.

If Shenzhen had not implemented mild intervention policies on January 23, 2020, the projected number of cumulative confirmed cases would be reach 1546 on February 3, 2020, which would be approximately 5.75 times of the actual number of cases.

The expected number of cumulative confirmed cases with corresponding 95% credible interval (CI) for different days of delay are summarized from January 29 to February 3, 2020 in Table 2. The full simulation results from January 19 to February 29, 2020 are presented in supplementary Appendix Figure A1. According to Table 2, the expected cumulative confirmed cases were 624 with the corresponding 95% CI (298,1551) for the 4-day delay and 676 with the corresponding 95% CI (303,1959) for the 5-day delay. Based on Figure A1, if the mild interventions were delayed for 4 or 5 days, the epidemic of COVID-19 in Shenzhen could be more severe than that in “synthetic Shenzhen” (1546 cumulative confirmed cases on February 3, 2020).

**Table 2.** *The likely outcome of intervention implemented delay in different days including the expected cumulative confirmed cases with corresponding 95% CI from January 29 to February 3, 2020*.

| Date | Actual | 0 day | 1 day | 2 days | 3 days | 4 days | 5 days |
| --- | --- | --- | --- | --- | --- | --- | --- |
| 01/29 | 86 | 92 (68,132) | 96 (69,146) | 98 (69,154) | 99 (69,159) | 99 (69,163) | 99 (69,165) |
| 01/30 | 110 | 126 (89,185) | 136 (91,216) | 141 (93,238) | 144 (93,258) | 145 (93,271) | 146 (93,281) |
| 01/31 | 170 | 163 (112,249) | 187 (122,309) | 202 (125,363) | 209 (126,407) | 213 (126,448) | 215 (126,490) |
| 02/01 | 196 | 201 (135,318) | 244 (155,415) | 281(164,522) | 300 (166,610) | 313 (167,716) | 319 (167,813) |
| 02/02 | 226 | 236 (154, 392) | 302 (188,518) | 370 (211,685) | 417 (225,862) | 450 (226,1098) | 469 (227,1294) |
| 02/03 | 269 | 269 (171, 458) | 355 (216, 616) | 456 (260,865) | 554 (288, 1175) | 624 (298,1551) | 676 (303,1959) |

## 4. DISCUSSION

We used the daily reported cumulative confirmed case data for 16 days since January 19, 2020, in Shenzhen and the corresponding data for 16 days since March 1, 2020 in 68 counties in the United States serving as a control group, i.e., the “synthetic Shenzhen”, those 68 United States counties were selected to match Shenzhen by latitude, population density and the first two components of 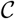 in (3) for the preintervention period. Those 68 United States counties did not implement systematic interventions during 16 days period.

By applying PCA to compress COVID-19 confirmed cases per 100,000 from the untreated units (counties in the United States), and combining the PCA with the latitude and population density in the estimation of weights through data assimilation, we were able to effectively estimate the model parameters based on the synthetic control method. In doing so, the latitude and population density as well as the principal components of 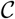 in (3) during pre-intervention period were considered as the characteristics of COVID-19.

The cases of COVID-19 per 100,000 in Shenzhen were significantly lower than those in “synthetic Shenzhen” after January 29, 2020. This indicated that the implementation of the mild intervention policies on January 23, 2020 in Shenzhen had a significant effect on controlling the epidemic of COVID-19. The reduction in the COVID-19 cases per 100,000 became larger and larger as time moved on (Figure 2).

The predicted cumulative confirmed cases (0-day delay) were very close to the observed cumulative confirmed cases of Shenzhen, indicating that our simulation could provide precise estimates[41]. Therefore, our simulated likely outcomes of delayed interventions were expected to be reliable. According to our simulation, there was little difference from January 23 to January 29, but the gap in the expected cumulative confirmed cases started increasing after January 29, 2020. This was consistent with the trend of the epidemic between Shenzhen and “synthetic Shenzhen” (Figure 2).

Therefore, it took about a week to see clear effects of interventions. If the mild interventions were not implemented, or were delayed by a week or longer, the epidemic of COVID-19 could have been severe. When the mild interventions were implemented early, the epidemic situation was significantly mitigated.

A novel step of our analysis strategy was to extend the use of SCM, which is a classical causal effect method and used it to create a synthetic reference population. Then, we introduced an epidemic compartmental SIHR model to evaluate the effectiveness of mild and early interventions in Shenzhen. Furthermore, the importance of the early interventions was revealed by simulating delays and assessing the consequences in the SIHR model. In conclusion, through the use of both SCM and SIHR models, the lessons were learned from the effect of timeliness of interventions to control the epidemic of COVID-19.

## Data Availability

Epidemic data were collected for daily cumulative confirmed cases of COVID-19 since January 19, 2020 in Shenzhen when the first confirmed case was reported. Its corresponding population, latitude and areas were collected in the statistical yearbook. Also, the daily cumulative confirmed cases of COVID-19 for each county of each state in the United States were downloaded and available online since March 1, 2020. The corresponding populations, areas, and latitudes were collected from the United States Census Bureau (USCB).

http://2019ncov.chinacdc.cn/2019-nCoV/index.html

http://2019ncov.chinacdc.cn/2019-nCoV/global.html

## APPENDIX A

### A.1 Effect of mild interventions

**Table A1.** *The pool of counties in the United States to synthesize Shenzhen with their corresponding population density, latitude and weights*.

| States | Counties | Latitude | Population density | Weights |
| --- | --- | --- | --- | --- |
| Guangdong | Shenzhen | 22.55 | 6670 |  |
| Florida | Broward | 26.19 | 570 | 0.423 |
| New York | New York | 40.78 | 18721 | 0.335 |
| California | Santa Clara | 37.22 | 571 | 0.123 |
| Florida | Pinellas | 27.90 | 619 | 0.032 |
| New York | Westchester | 41.15 | 747 | 0.025 |
| Florida | Orange | 28.51 | 536 | 0.016 |
| Florida | Seminole | 28.69 | 528 | 0.014 |
| Texas | Bexar | 29.45 | 616 | 0.008 |
| Texas | Harris | 29.86 | 1024 | 0.007 |
| Georgia | Fulton | 33.79 | 769 | 0.002 |
| California | Los Angeles | 34.20 | 816 | 0.001 |
| California | Orange | 33.68 | 1293 | 0.001 |
| Georgia | Clayton | 33.55 | 782 | 0.001 |
| Georgia | Cobb | 33.94 | 852 | 0.001 |
| Georgia | DeKalb | 33.77 | 1081 | 0.001 |
| Georgia | Gwinnett | 33.96 | 828 | 0.001 |
| Texas | Dallas | 32.77 | 1120 | 0.001 |
| Texas | Tarrant | 32.77 | 900 | 0.001 |

**Figure A1.**
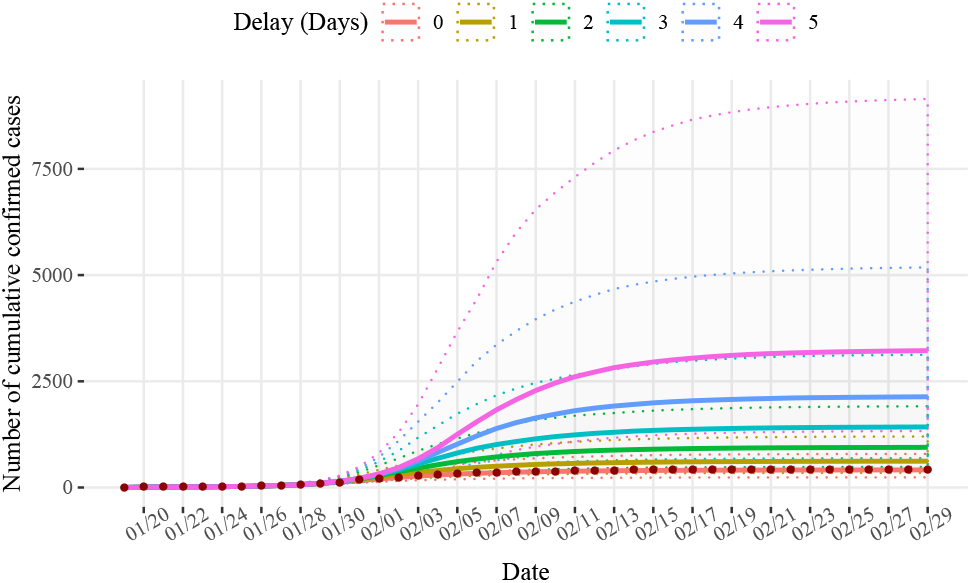
*The expected number of cumulative confirmed cases with corresponding confidence intervals for mild interventions if they were delayed by different days from January 19 to February 29, 2020. The points represent the actual cumulative confirmed cases of Shenzhen*.

### A.2 Delay effect of mild interventions

The dynamic system of compartments *S*, *I*, *H*, *R* are as follows:

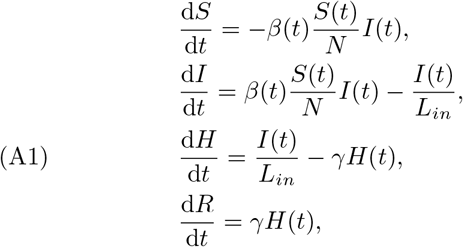

where (*S*(*t*), *I*(*t*), *H*(*t*), *R*(*t*)) are the numbers of corresponding compartments at the time *t*. Given initial (*S*(*t*_0_), *I*(*t*_0_), *H*(*t*_0_), *R*(*t*_0_)) and the parameters (*β*(*t*), *L_in_*, γ), we can simulate the (*S*(*t*_1_), *I*(*t*_1_),*H*(*t*_1_),*R*(*t*_1_)), *t*_1_ > *t*_0_ by the deterministic dynamic system (A1). For simplicity, let *β*(*t*) ≡ *β*(*k*), if *k* ≤ *t* < *k* + 1.

### A.3 Online Materials

The data and R files supporting the conclusions of this study are available in the https://github.com/tingT0929/The-timing-and-effectiveness-of-implementing-mild-interventions-of-COVID-19

## ACKNOWLEDGEMENTS

We would like to thank all individuals who are collecting epidemiological data of the COVID-19 outbreak around the world.

Ting Tian

School of Mathematics Sun Yat-Sen University Guangzhou, 510275 China

Wenxiang Luo School of Mathematics Sun Yat-Sen University Guangzhou, GD 510275 China

Jianbin Tan School of Mathematics Sun Yat-Sen University Guangzhou, GD 510275 China

Yukang Jiang School of Mathematics Sun Yat-Sen University Guangzhou, GD 510275 China

Minqiong Chen School of Mathematics Sun Yat-Sen University Guangzhou, GD 510275 China

Wenliang Pan

School of Mathematics Sun Yat-Sen University Guangzhou, GD 510275 China

Songpan Yang School of Mathematics Sun Yat-Sen University Guangzhou, GD 510275 China

Jiashu Zhao School of Mathematics Sun Yat-Sen University Guangzhou, GD 510275 China

Xueqin Wang School of Statistics

Capital University of Economics and Business Beijing, 100070 China

Heping Zhang

School of Public Health Yale University New Haven, CT 06520-8034 the United States

